# Prognostic Value of Angiography-derived Microcirculatory Resistance in Patients undergoing Rotational Atherectomy

**DOI:** 10.1101/2023.11.13.23298493

**Authors:** Xi Zhang, Qing Jin, Jiaji He, Tao Zhao, Guiping He, Qiang Xue, Xuefeng Guang

## Abstract

**BACKGROUND:** Rotational atherectomy (RA) is predominantly employed in the treatment of severe calcification lesions in patients with coronary atherosclerotic heart disease (CAD). Studies focusing on the assessment of postoperative microvascular dysfunction (CMD) after RA and related prognosis are scarce.

**AIMS:** we attempted to investigate the predictive significance of coronary angiography-derived microcirculatory resistance (AMR) in patients with coronary RA.

**METHODS:** This retrospective study analyzed the data from 114 patients who were successfully treated between January 2019 and September 2022. Coronary microcirculatory function after RA was assessed using AMR. Patients were categorized into CMD and non-CMD groups depending on a postoperative AMR of ≥2.5 mmHg-s/cm. Patients were followed up for postoperative major adverse cardiovascular events (MACE).

**RESULTS:** We analyzed the data from 114 patients, and post-RA, the mean AMR, mean QFR, and the percentage of CMDs were significantly higher compared to those before RA. MACE occurred in 14 (12.3%) patients after a year of follow-up. A higher proportion of patients in the MACE group showed post-RA AMR of ≥2.5 mmHg-s/cm (57.1% vs. 27.0%, P=0.048). Cox regression analysis showed that AMR ≥2.5 mmHg-s/cm (HR=3.86, 95%CI. 1.28-11.63, P=0.016) and renal insufficiency (HR=9.92, 95%CI: 2.06-47.83, P=0.004) were independent predictors of MACE. Logistic regression analyses showed the length of the RA operative area and diabetes mellitus (DM) were related to post-RA CMD.

**CONCLUSION:** In patients with CAD treated with RA, AMR ≥2.5 mmHg-s/cm independently predicted post-RA MACE; furthermore, the operative length of RA and the comorbid DM were associated with CMD following RA.

## Background

Coronary atherosclerotic heart disease (CAD), with a frequent incidence, poses a serious threat to human health worldwide,[1, 2] and nearly 30% of these cases are associated with moderate or severe coronary artery calcification.[3–5] Calcified lesions pose many challenges to percutaneous coronary intervention (PCI), such as intraoperative balloon dilatation stuck, difficult stent passage or incomplete stent expansion, and increased risk of postprocedural stent restenosis and thrombosis, resulting in an enhanced risk of ischemic events which seriously affect the long-term prognosis.[4, 6, 7] Rotational atherectomy (RA) is increasingly being recognized as improving the surgical success rate by breaking down calcified plaques through high-speed rotation of the rotary head.[8, 9] However, its use may lead to vascular and microcirculatory damage, further exacerbating myocardial ischemia and leading to the deterioration of cardiac function.[10]

At present, clinical assessment of microcirculation mainly encompasses coronary flow reserve (CFR), index of microvascular resistance (IMR), etc., which have not been widely adopted owing to limitations such as complex operative processes and unstable measurement. Angiographic microvascular resistance (AMR) from coronary angiography does not require the use of pressure guidewire and vasodilator drugs, is simple to operate, and can achieve the measurement of microcirculatory resistance within 1–2 min. It has the advantages of being economical and simple. Relevant studies have shown AMR’s high accuracy and suitability for clinical use.[11]

Studies on the assessment of post-RA coronary microvascular dysfunction (CMD) and related prognoses are scanty. To address this, we designed a retrospective, single-center clinical trial to assess the effect of RA on postoperative microcirculatory function in patients with severe calcification using AMR and determine its correlation with major adverse cardiovascular events (MACE) after RA.

### Study design

This was a retrospective, single-center study that complied with medical ethics and was approved by the Ethics Committee of our hospital (YAXLL-AF-SC-021/01). All procedures performed on patients followed the Helsinki Declaration

### Inclusion criteria

Consecutively enrolled patients with severe calcification of coronary arteries who received intraoperative RA treatment between January 2019 and September 2022 in the Yan’an Hospital, Kunming, China. The inclusion criteria were (1) age ≥18 years and (2) the presence of severe calcification in at least one vessel that was successfully treated with RA. Severe calcification was defined either as coronary angiography (CAG) showing a dense shadow that traveled along the vessel wall and was visualized during the cardiac cycle, or intravascular ultrasound (IVUS) showing a hyperechoic plaque with posterior acoustic shadows at > 180°.

Exclusion criteria in the clinical settings were (1) patients who did not receive treatment with RA, (2) unavailability of follow-up data, and (3) acute myocardial infarction within 7 days of RA. Angiography exclusion criteria were (1) missing preoperative or postoperative contrast and (2) AMR analysis was not possible because of the inability to detect vessel borders or poorly filled contrast, excessive overlap of stenotic segments, or severely tortuous lesions in the target vessel. All interventions performed for patients were in line with the guidelines outlined in the Declaration of Helsinki. Informed consent of patients was waived-off because all data in this study were retrospectively collected.

### Procedure of RA

All RA surgeries were performed following the standard therapeutic procedures outlined in the extant guidelines. The rotary mill head diameter ranged from 1.25 to 2.0 mm and speed was maintained between 13 and 22 × 10^4^ rpm.[8, 12, 13] Intraoperative irrigation with heparinized saline was routinely performed to reduce the incidence of decreased blood flow. Intraoperative surgical procedures such as IVUS evaluation, cutting balloon dilatation, or drug-eluting stent placement were decided by the operator according to the lesional characteristics. The surgery was considered successful when CAG suggested that the residual stenosis of the vessel was small, i.e., ≤30% and the TIMI flow was graded III. For intraoperative complications such as decreased blood pressure, hematoma, and entrapment, patients received standard treatment and were documented in the RA surgical management system. All patients received dual antiplatelet therapy and secondary prevention of coronary heart disease after surgery was exercised. Preoperative and postoperative imaging data were preserved for calculating Murray’s law-based quantitative flow ratio (μQFR) and AMR.

### AMR and QFR measurements

The CAG image meeting the criterion was imported in AngioPlus software (Pulse Medical Technology, Shanghai, China). μQFR and AMR calculation methods have been previously published. Briefly, coronary arteries were revascularized using Murray’s law, and μQFR was derived along with simulated congestive state flow velocities.[14] Distal coronary pressure (Pd) was obtained based on the pressure drop, and AMR = Pd / simulated hyperemic flow velocity (*Velocity*hyp) was calculated as shown in Fig. 2. [15] Patients with an AMR ≥2.5 mmHg-s/cm were defined as those with CMD.

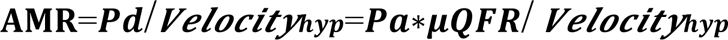

AMR and QFR were measured by two specialists blinded to the patient’s clinical characteristics and information related to RA surgery and clinical outcomes. In case of any contradiction such as inconsistency in the results, the data were analyzed by a third specialist, and agreement was reached after discussion. ΔAMR was calculated as post AMR-pre AMR.

AMR and QFR were measured by two specialists blinded to the patient’s clinical characteristics and information related to RA surgery and clinical outcomes. In case of any contradiction such as inconsistency in the results, the data were analyzed by a third specialist, and agreement was reached after discussion. ΔAMR was calculated as post AMR-pre AMR.

### Follow-up and clinical outcomes

Patients meeting the inclusion criteria were included and their clinical data were collected. Follow-up visits for patients who were discharged from the hospital were completed periodically through outpatient or inpatient visits or by telephone. To avoid confounding factors, all data were collected based on information recorded in the hospital’s electronic system. Clinical endpoints in this study were defined as MACE (all-cause death, nonfatal myocardial infarction, revascularization, and angina-related rehospitalization).

### Statistical methods

Histograms were used to assess data distribution. Normally distributed continuous variables are expressed as mean ± standard deviation and non-normally distributed data as median and interquartile range. Categorical variables are expressed as frequencies and percentages. Comparison between baseline and follow-up normally distributed data was performed using t-tests, while non-normally distributed data were compared using Wilcoxon signed-rank tests.

Kaplan-Meier analysis of event-free survival (EFS) allowed the assessment of possible risk factors (sex, age, smoking, CMD, etc.), and their significance was tested by a log-rank test. Univariate COX analysis with *P*<0.10 was considered statistically significant and included as an independent variable in the COX regression model. Likelihood ratios (LRs) were used to confirm reasonably and statistically significant interactions. The predictive effect of CMD on MACE was assessed using the COX regression model. Univariate and subsequent multivariate logistic regression analyses were used to determine associations between patient characteristics, surgical factors related to RA, and CMD after RA. Differences were considered statistically significant for *P*<0.05. Statistical analysis was performed using SPSS Statistics, version 26 (IBM Inc, Armonk, NY, USA).

## Results

### 1 Patients’ baseline characteristics

In total, 121 patients who met the inclusion criteria were included. As shown in Figure 1, follow-up was completed for 114 patients (94.2%). The clinical and angiographic characteristics of the included patients are shown in Table 1. The mean age was 66.1 years and 56 (49.1%) were male. Seventy-one (62.3%) suffered from hypertension, 46 (40.4%) had dyslipidemia, 48 (42.1%) had DM; 15 (13.2%) had previous myocardial infarction and six (5.3%) had PCI history. All patients showed multiple coronary artery lesions with 55 patients (48.2%) having three lesions. Among vessels subjected to RA, the distribution was 79 (69.3%) patients with left anterior descending, 6 (5.3%) with left echogenic, and 24 (21.1%) with right coronary vessels.

**Fig. 1.**
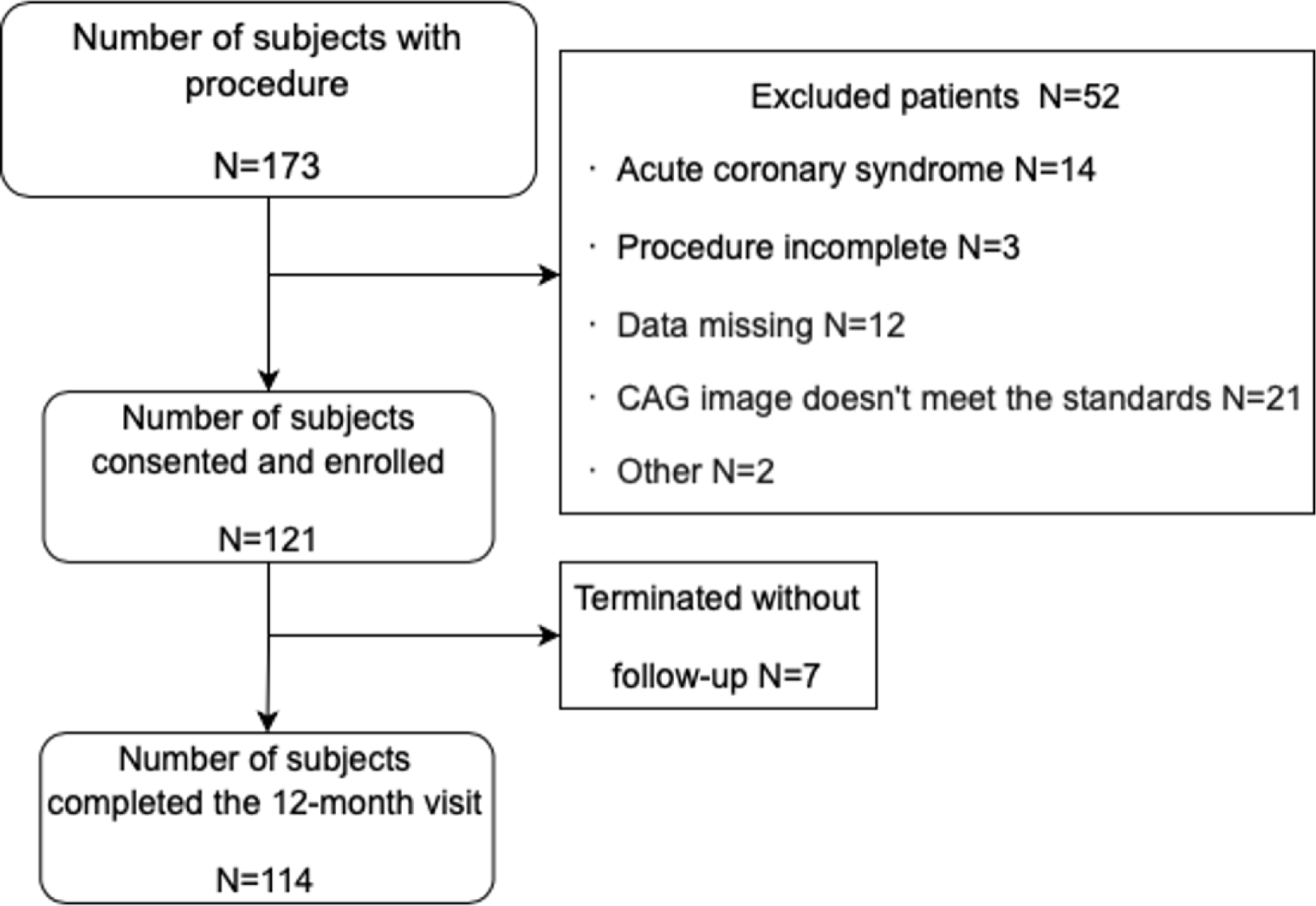
Flow chart of enrolled patients.

**Fig. 2.**
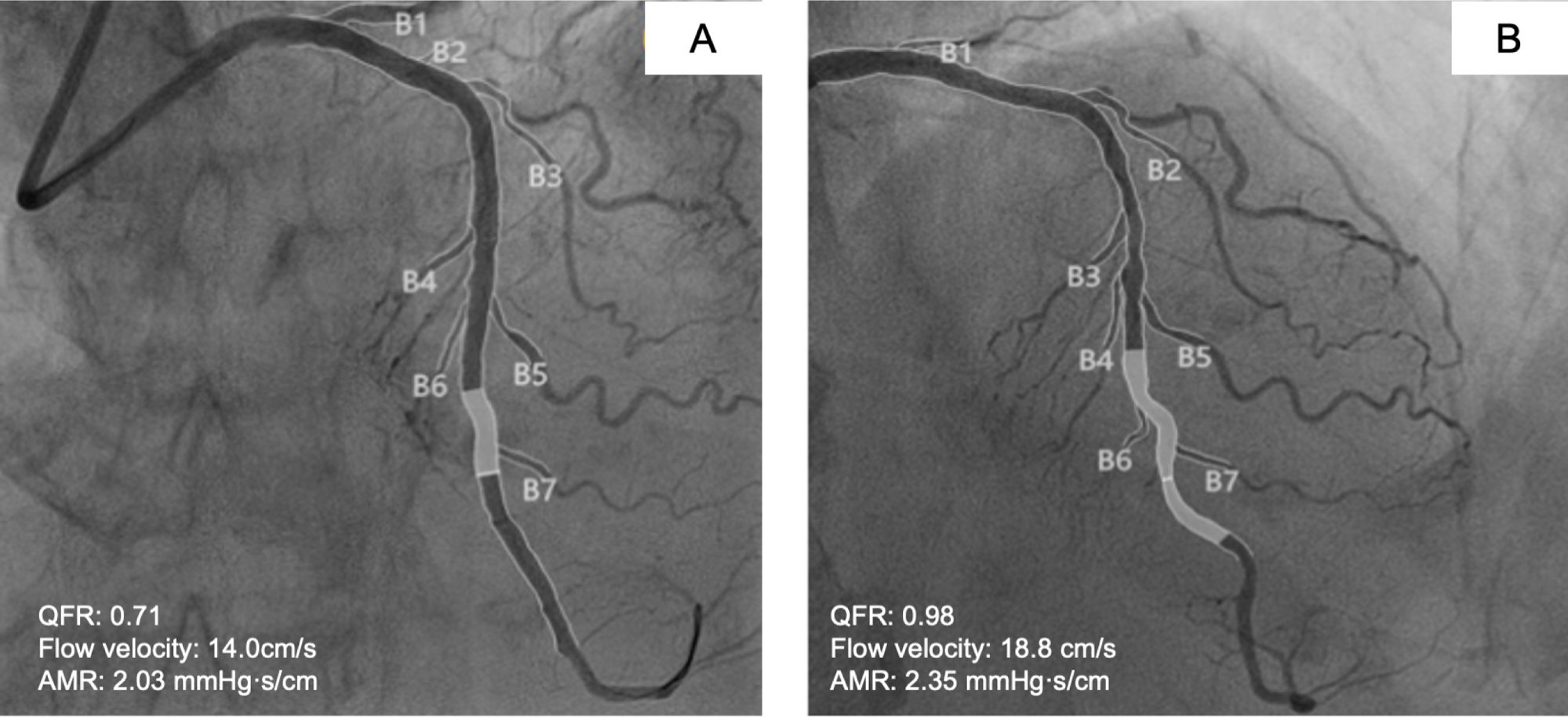
Examples of QFR and AMR analysis. CAG showed a stenosis in the left anterior descending, and QFR was calculated as 0.71, flow velocity was 14.0 *cm/s,* AMR was calculated as 2.03 mmHg·sfcm (A). After RA, QFR was calculated as 0.98, flow velocity was 18.8 *cm/s,* and AMR was calculated as 2.35 mmHg·sfcm (B). RA, rotational atherectomy;QFR, quantitative flow ratio; AMR, angiographic microvascular resistance; CAG, coronary angiography.

**Table1.**
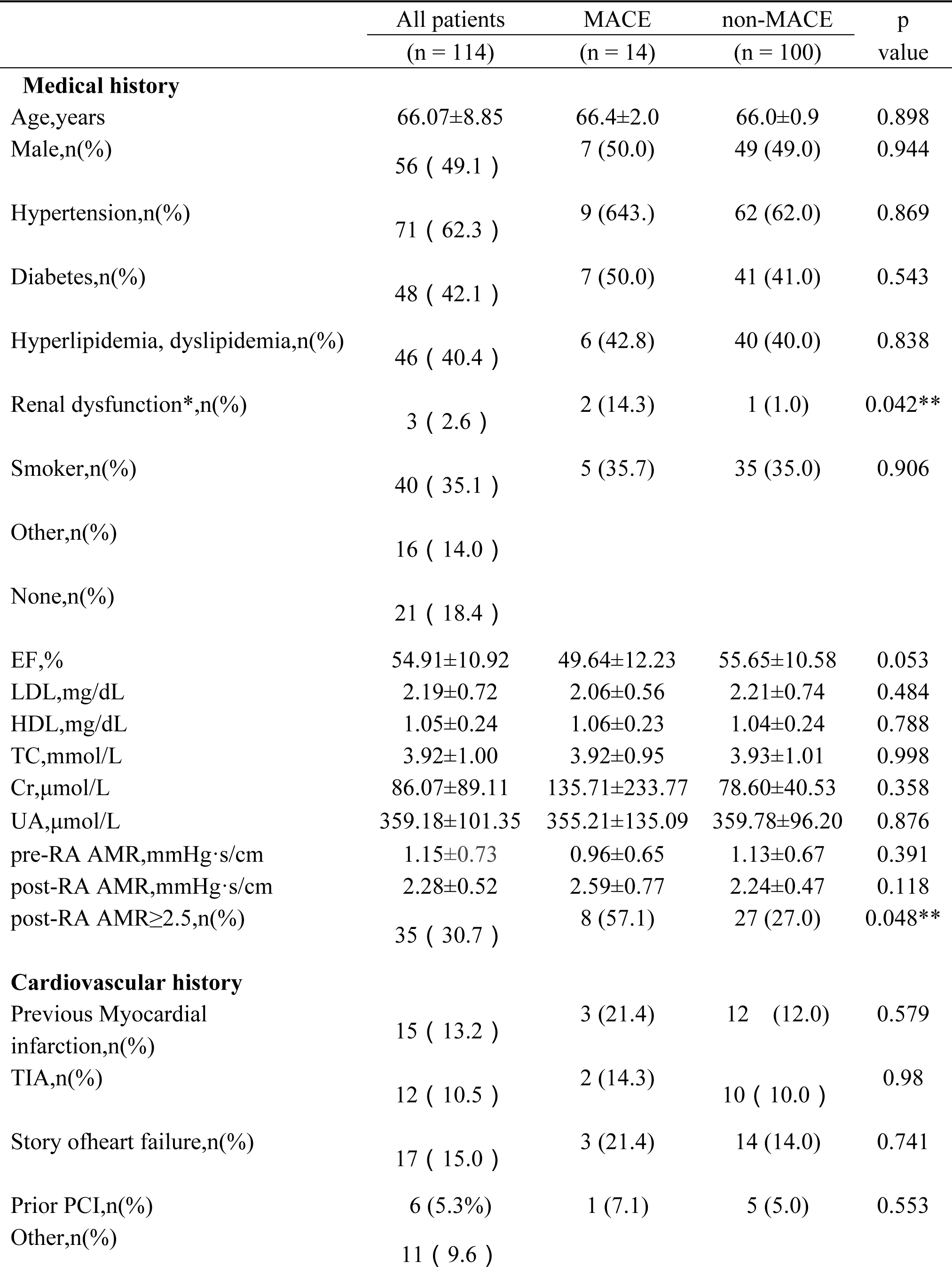
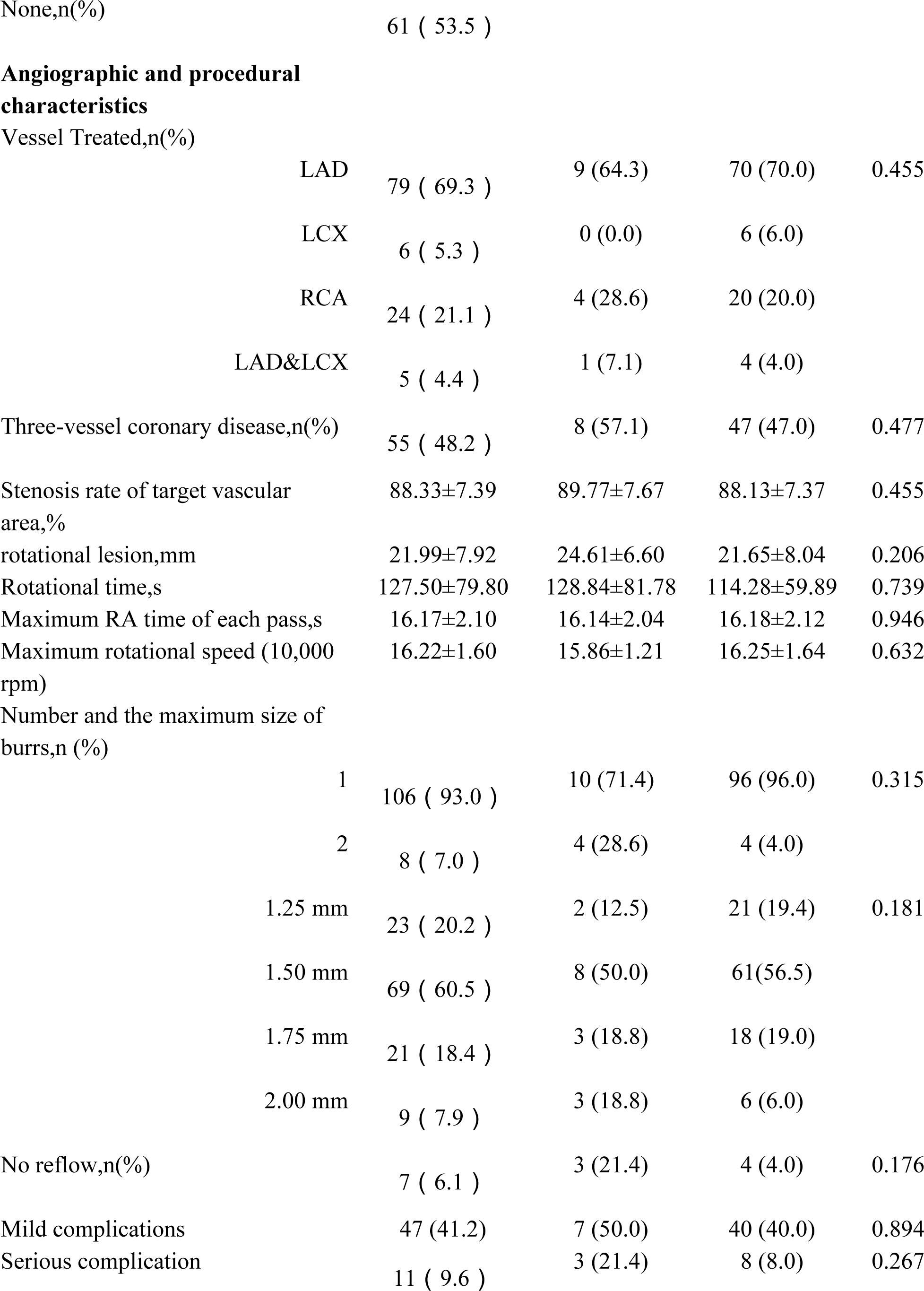
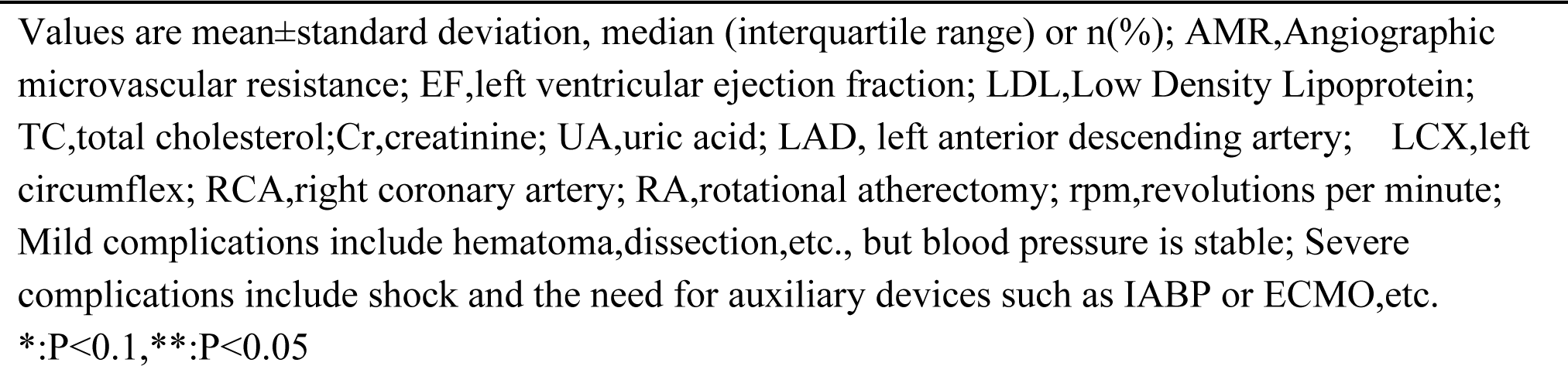
Baseline characteristics of the study population.

|  | All patients<br>(n = 114) | MACE<br>(n = 14) | non-MACE<br>(n = 100) | p<br>value |
| --- | --- | --- | --- | --- |
| **Medical history** |  |  |  |  |
| Age,years | 66.07±8.85 | 66.4±2.0 | 66.0±0.9 | 0.898 |
| Male,n(%) | 56 ( 49.1 ) | 7 (50.0) | 49 (49.0) | 0.944 |
| Hypertension,n(%) | 71 ( 62.3 ) | 9 (64.3) | 62 (62.0) | 0.869 |
| Diabetes,n(%) | 48 ( 42.1 ) | 7 (50.0) | 41 (41.0) | 0.543 |
| Hyperlipidemia, dyslipidemia,n(%) | 46 ( 40.4 ) | 6 (42.8) | 40 (40.0) | 0.838 |
| Renal dysfunction*,n(%) | 3 ( 2.6 ) | 2 (14.3) | 1 (1.0) | 0.042** |
| Smoker,n(%) | 40 ( 35.1 ) | 5 (35.7) | 35 (35.0) | 0.906 |
| Other,n(%) | 16 ( 14.0 ) |  |  |  |
| None,n(%) | 21 ( 18.4 ) |  |  |  |
| EF,% | 54.91±10.92 | 49.64±12.23 | 55.65±10.58 | 0.053 |
| LDL,mg/dL | 2.19±0.72 | 2.06±0.56 | 2.21±0.74 | 0.484 |
| HDL,mg/dL | 1.05±0.24 | 1.06±0.23 | 1.04±0.24 | 0.788 |
| TC,mmol/L | 3.92±1.00 | 3.92±0.95 | 3.93±1.01 | 0.998 |
| Cr,μmol/L | 86.07±89.11 | 135.71±233.77 | 78.60±40.53 | 0.358 |
| UA,μmol/L | 359.18±101.35 | 355.21±135.09 | 359.78±96.20 | 0.876 |
| pre-RA AMR,mmHg·s/cm | 1.15±0.73 | 0.96±0.65 | 1.13±0.67 | 0.391 |
| post-RA AMR,mmHg·s/cm | 2.28±0.52 | 2.59±0.77 | 2.24±0.47 | 0.118 |
| post-RA AMR≥2.5,n(%) | 35 ( 30.7 ) | 8 (57.1) | 27 (27.0) | 0.048** |
| **Cardiovascular history** |  |  |  |  |
| Previous Myocardial infarction,n(%) | 15 ( 13.2 ) | 3 (21.4) | 12 (12.0) | 0.579 |
| TIA,n(%) | 12 ( 10.5 ) | 2 (14.3) | 10 ( 10.0 ) | 0.98 |
| Story of heart failure,n(%) | 17 ( 15.0 ) | 3 (21.4) | 14 (14.0) | 0.741 |
| Prior PCI,n(%) | 6 (5.3%) | 1 (7.1) | 5 (5.0) | 0.553 |
| Other,n(%) | 11 ( 9.6 ) |  |  |  |

|  |  |  |  |  |  |
| --- | --- | --- | --- | --- | --- |
| None,n(%) |  | 61 ( 53.5 ) |  |  |  |
| **Angiographic and procedural characteristics** |  |  |  |  |  |
| Vessel Treated,n(%) |  |  |  |  |  |
|  | LAD | 79 ( 69.3 ) | 9 (64.3) | 70 (70.0) | 0.455 |
|  | LCX | 6 ( 5.3 ) | 0 (0.0) | 6 (6.0) |  |
|  | RCA | 24 ( 21.1 ) | 4 (28.6) | 20 (20.0) |  |
|  | LAD&LCX | 5 ( 4.4 ) | 1 (7.1) | 4 (4.0) |  |
| Three-vessel coronary disease,n(%) |  | 55 ( 48.2 ) | 8 (57.1) | 47 (47.0) | 0.477 |
| Stenosis rate of target vascular area,% |  | 88.33±7.39 | 89.77±7.67 | 88.13±7.37 | 0.455 |
| rotational lesion,mm |  | 21.99±7.92 | 24.61±6.60 | 21.65±8.04 | 0.206 |
| Rotational time,s |  | 127.50±79.80 | 128.84±81.78 | 114.28±59.89 | 0.739 |
| Maximum RA time of each pass,s |  | 16.17±2.10 | 16.14±2.04 | 16.18±2.12 | 0.946 |
| Maximum rotational speed (10,000 rpm) |  | 16.22±1.60 | 15.86±1.21 | 16.25±1.64 | 0.632 |
| Number and the maximum size of burrs,n (%) |  |  |  |  |  |
|  | 1 | 106 ( 93.0 ) | 10 (71.4) | 96 (96.0) | 0.315 |
|  | 2 | 8 ( 7.0 ) | 4 (28.6) | 4 (4.0) |  |
|  | 1.25 mm | 23 ( 20.2 ) | 2 (12.5) | 21 (19.4) | 0.181 |
|  | 1.50 mm | 69 ( 60.5 ) | 8 (50.0) | 61(56.5) |  |
|  | 1.75 mm | 21 ( 18.4 ) | 3 (18.8) | 18 (19.0) |  |
|  | 2.00 mm | 9 ( 7.9 ) | 3 (18.8) | 6 (6.0) |  |
| No reflow,n(%) |  | 7 ( 6.1 ) | 3 (21.4) | 4 (4.0) | 0.176 |
| Mild complications |  | 47 (41.2) | 7 (50.0) | 40 (40.0) | 0.894 |
| Serious complication |  | 11 ( 9.6 ) | 3 (21.4) | 8 (8.0) | 0.267 |

Values are mean±standard deviation, median (interquartile range) or n(%); AMR,Angiographic microvascular resistance; EF,left ventricular ejection fraction; LDL,Low Density Lipoprotein; TC,total cholesterol;Cr,creatinine; UA,uric acid; LAD, left anterior descending artery; LCX,left circumflex; RCA,right coronary artery; RA,rotational atherectomy; rpm,revolutions per minute; Mild complications include hematoma,dissection,etc., but blood pressure is stable; Severe complications include shock and the need for auxiliary devices such as IABP or ECMO,etc. \*:P<0.1, \*\*:P<0.05

### 2 QFR and AMR results

The mean post-AR AMR (2.28±0.52 vs. 1.15±0.73, *P*<0.0001) and QFR (0.92 ±0.01 vs. 0.41±0.02, *P*<0.0001) were significantly higher than the values before RA, as shown in Figure 3. Patients were categorized into CMD and non-CMD groups based on the postoperative AMR, and the value in the former was significantly higher than that in the latter (2.89±0.43 vs. 2.02±0.38, *P*<0.0001). Thirty-five patients (30.7%) suffered CMD after RA, significantly higher than the pre-RA CMD rates (*P*<0.0001, Figure 4A).

**Fig. 3.**
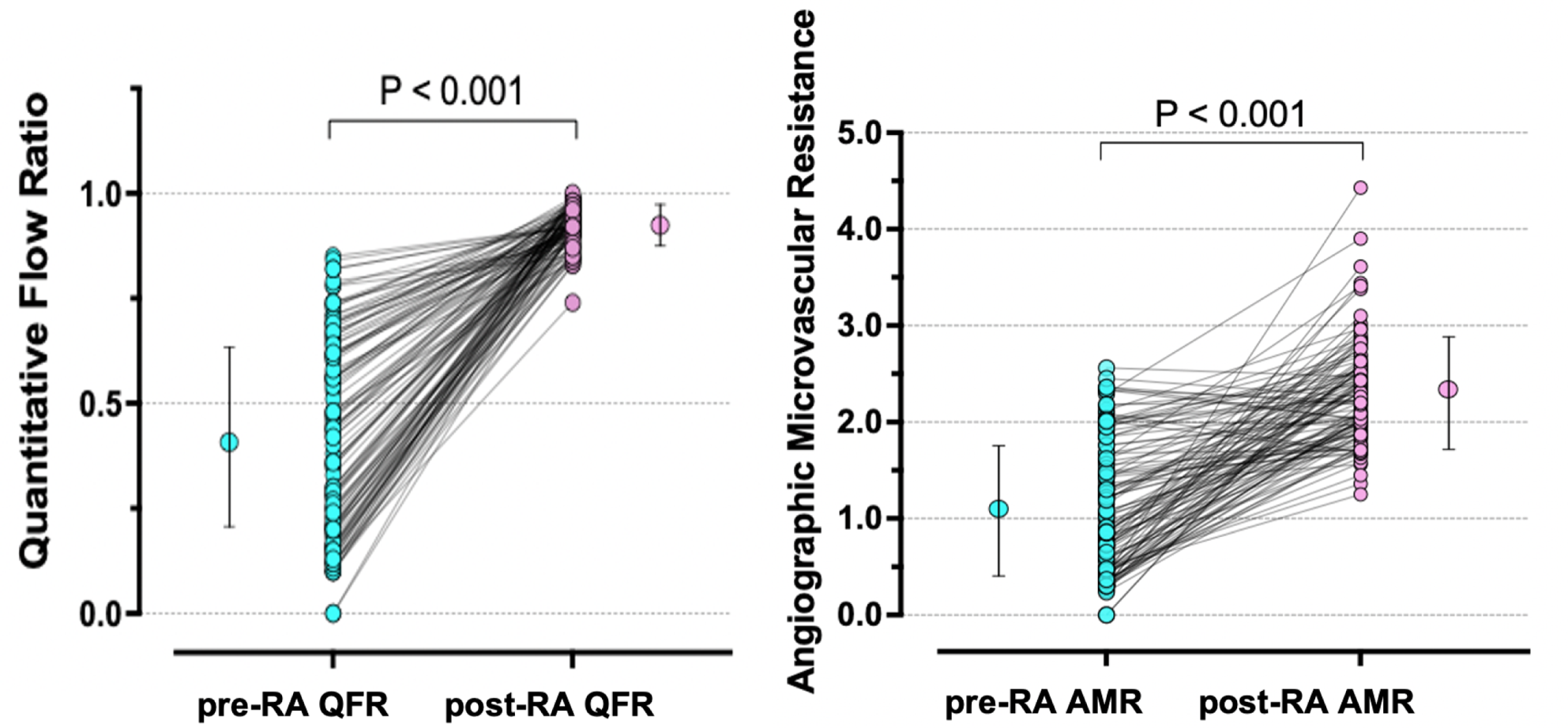
Temporal change of Quantitative flow ratio (QFR) and Angiographic microvascular resistance (AMR) in the target vessels. The QFR and AMR significantly improved in patients from pre-RA to post-RA. QFR, Quantitative flow ratio; AMR, Angiographic microvascular resistance; RA, rotational atherectomy.

**Fig. 4.**
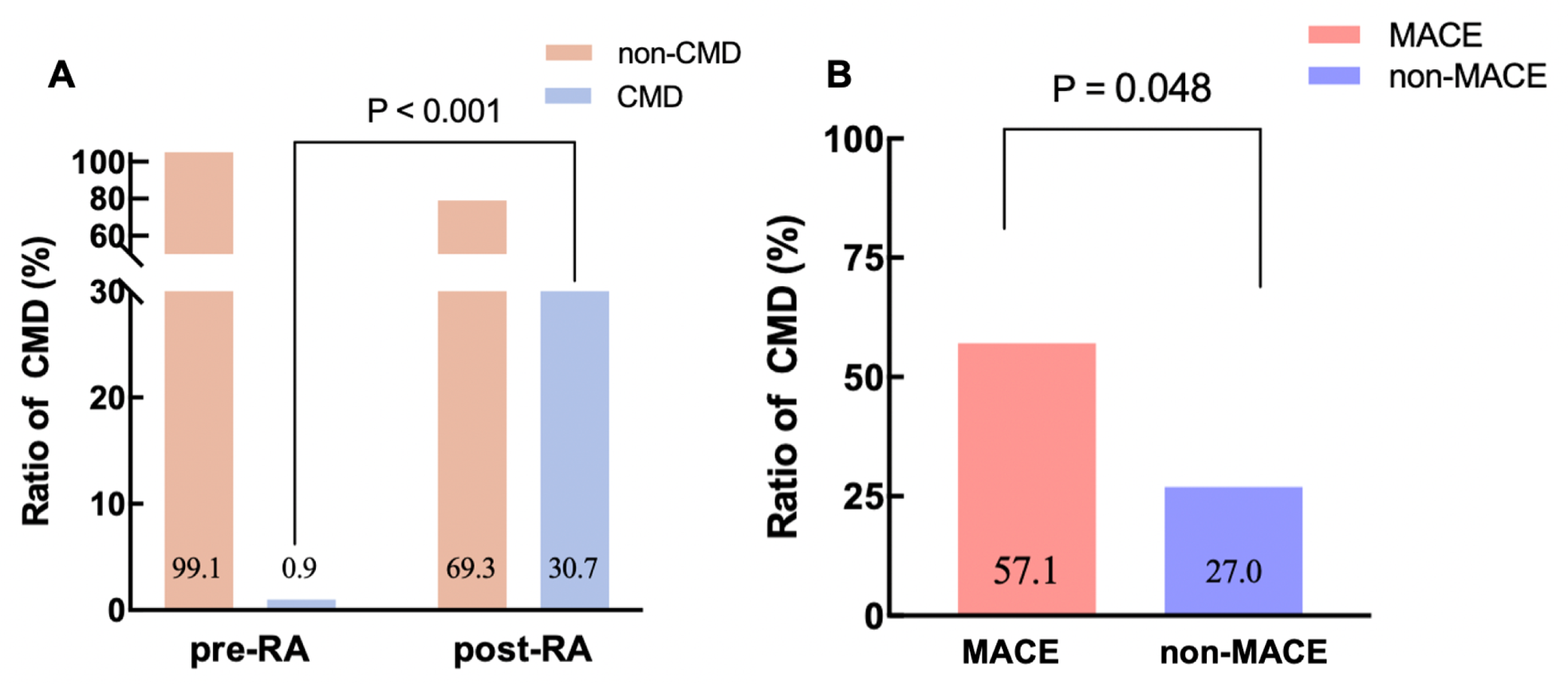
Coronary microvascular dysfunction(CMD) ratio of patients pre-RA vs. post-RA (A), and MACE vs. non-MACE. (A) The ratio of patients who CMD in post-RA higher than pre-RA. (B) The ratio of patients who CMD in MACE higher than non-MACE.AMR. AMR, Angiographic microvascular resistance; RA, rotational atherectomy. CMD(AMR 2.5); non-CMD (AMR<2.5); MACE: all-cause death, nonfatal myocardial infarction, revascularization, angina-related rehospitalization.

### 3 Correlation between AMR and MACE following RA and the main prognostic predictors for patients with RA

Fourteen patients (12.3%) showed MACE events during the follow-up. Two died of cardiac causes; six underwent revascularization; two suffered from stroke, and two died of non-cardiac causes. The patients were categorized into MACE and non-MACE groups based on the occurrence of MACE events.

There was no significant difference in preoperative AMR in patients who developed MACE compared to those who did not (0.96±0.65 vs. 1.13±0.67, *P*=0.391). However, there was a trend toward higher postoperative AMR in patients in the MACE group (2.59±0.77 vs. 2.24±0.47, *P*=0.118, Table 1). The proportion of patients with CMD was higher in the MACE group (57.1% vs 27.0%, *P*=0.048, Figure 4B).

Kaplan-Meier curve analysis showed a significantly higher incidence of MACE in postoperative AMR-defined CMD compared with non-CMD (22.8% vs. 7.6%; HR=3.17; 95% CI:1.10–9.14; log-rank *P*=0.023) among patients as shown in Figure 5. Univariate and multivariate Cox regression results of MACE predictors are shown in Table 2. Post-RA AMR ≥2.5 and renal insufficiency were independently associated with MACE, evidenced by the results of univariate Cox regression. After adjusting for confounders, post-RA AMR≥2.5 (HR=3.86,95%CI:1.28-11.63, *P*=0.016) and renal insufficiency (HR=9.92,95%CI:2.06-47.83, *P*=0.004) remained statistically significant in the multivariate Cox regression model as an independent predictor of primary outcome in patients with RA (post-refers to ‘after RA operation’; pre-RA indicates ‘before RA operation’).

**Fig. 5.**
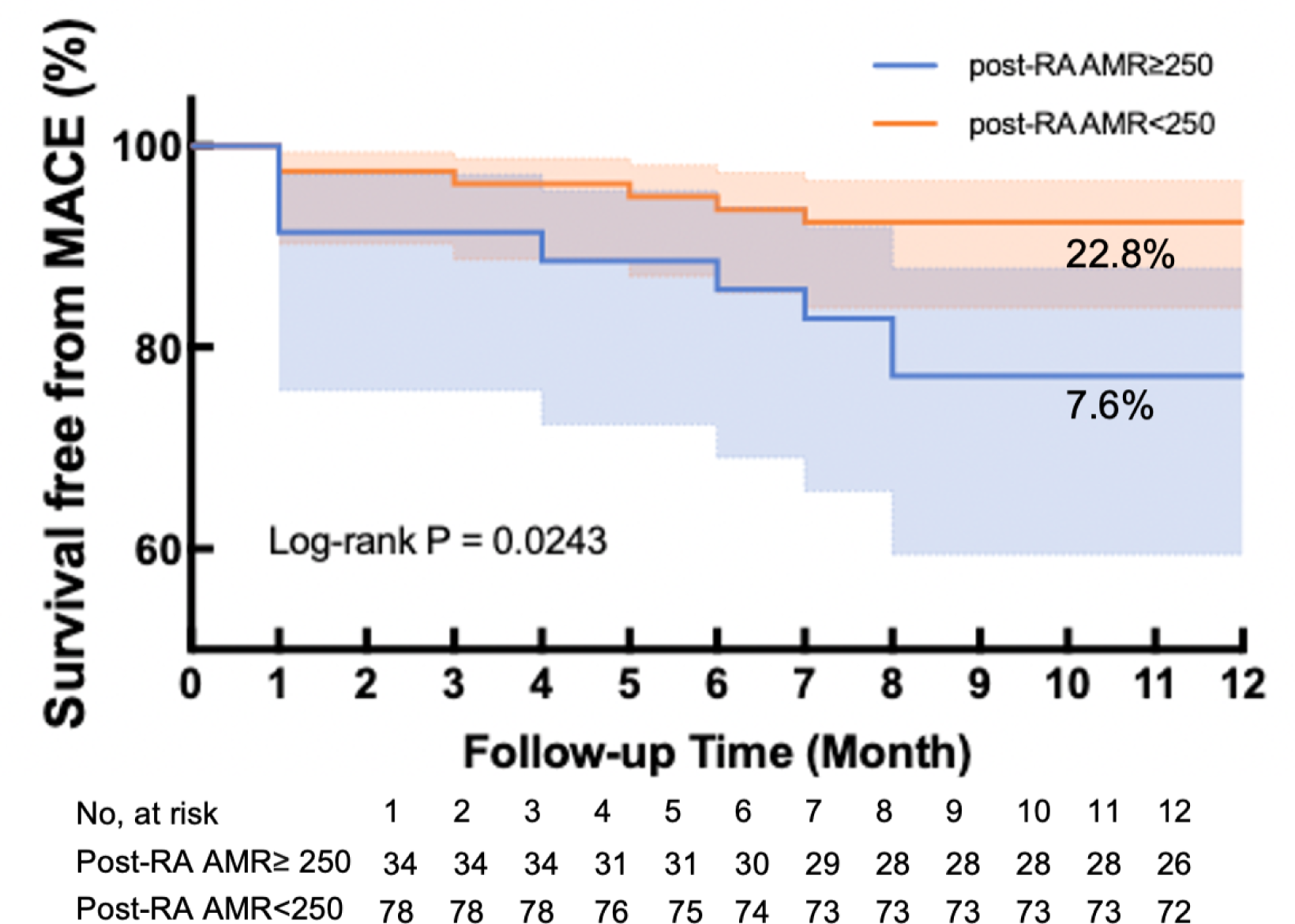
Kaplan-Meier curves for freedom from MACE stratified by post-RA AMR. AMR, Angiographic microvascular resistance; RA, rotational atherectomy. MACE: all-cause death, nonfatal myocardial infarction, revascularization, angina-related rehospitalization

**Table2.**
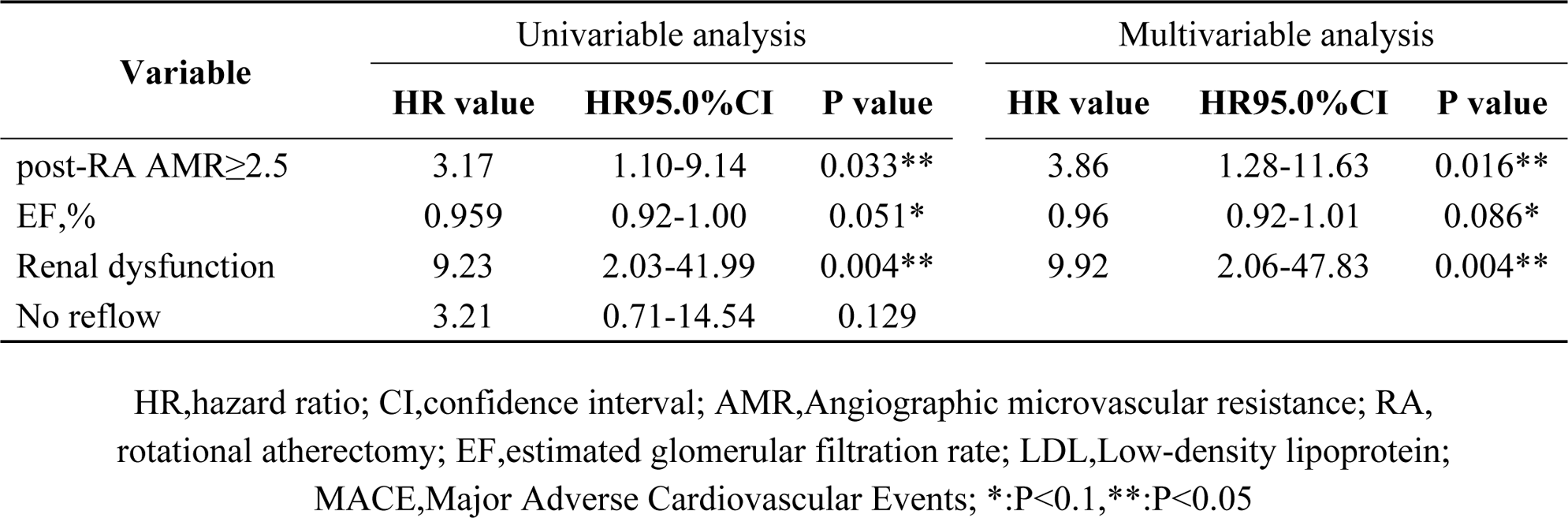
Predictors of the MACE in patients who underwent RA.

| Variable | Univariable analysis |  |  | Multivariable analysis |  |  |
| --- | --- | --- | --- | --- | --- | --- |
|  | HR value | HR95.0%CI | P value | HR value | HR95.0%CI | P value |
| post-RA AMR≥2.5 | 3.17 | 1.10-9.14 | 0.033** | 3.86 | 1.28-11.63 | 0.016** |
| EF,% | 0.959 | 0.92-1.00 | 0.051* | 0.96 | 0.92-1.01 | 0.086* |
| Renal dysfunction | 9.23 | 2.03-41.99 | 0.004** | 9.92 | 2.06-47.83 | 0.004** |
| No reflow | 3.21 | 0.71-14.54 | 0.129 |  |  |  |

HR,hazard ratio; CI,confidence interval; AMR,Angiographic microvascular resistance; RA, rotational atherectomy; EF,estimated glomerular filtration rate; LDL,Low-density lipoprotein; MACE,Major Adverse Cardiovascular Events; \*:P<0.1, \*\*:P<0.05

### 4 Predictors of postoperative CMD in patients with RA

Table 3 shows the predictors of postoperative CMD in RA obtained by using univariate and multivariate logistic regression models. The significant predictors in the univariate analysis included age, hypertension, DM, previous myocardial infarction, creatinine, number of variable vessels, lesion length, three lesion lesions, maximum diameter and number of rotary grinders, maximum duration of a single RA, total duration of RA, and complications. Multivariate analysis showed that independent predictors of postoperative CMD were lesion length (OR=2.03, 95% CI: 1.16–3.59, *P*=0.014) and diabetes (OR=2.66, 95% CI: 1.12–6.31, *P*=0.027).

**Table3.**
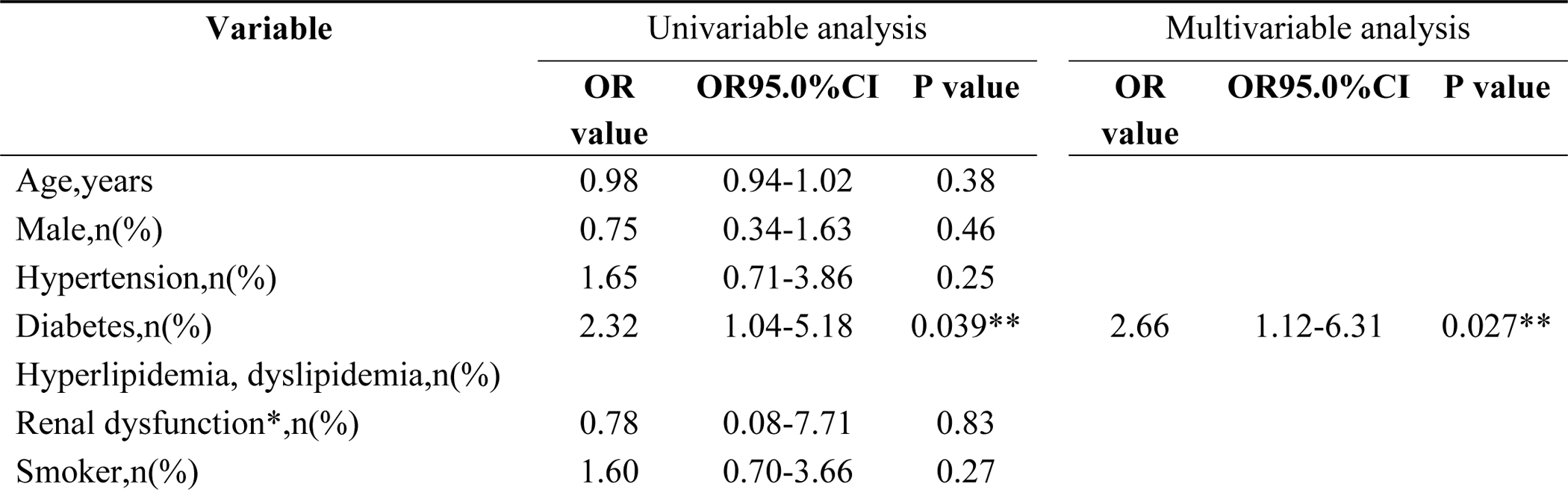
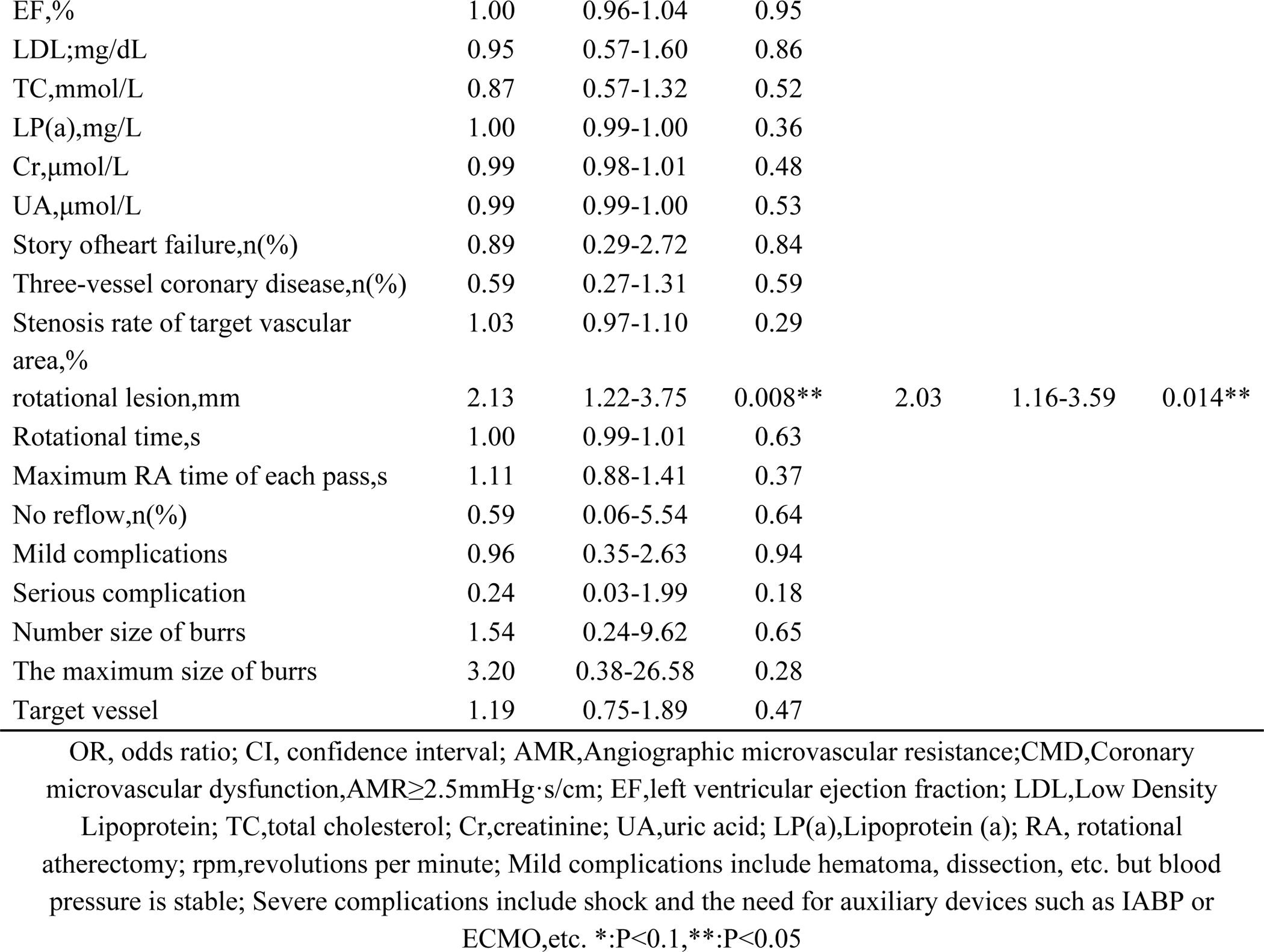
Independent predictors of CMD.

| Variable | Univariable analysis |  |  | Multivariable analysis |  |  |
| --- | --- | --- | --- | --- | --- | --- |
|  | OR<br>value | OR95.0%CI | P value | OR<br>value | OR95.0%CI | P value |
| Age,years | 0.98 | 0.94-1.02 | 0.38 |  |  |  |
| Male,n(%) | 0.75 | 0.34-1.63 | 0.46 |  |  |  |
| Hypertension,n(%) | 1.65 | 0.71-3.86 | 0.25 |  |  |  |
| Diabetes,n(%) | 2.32 | 1.04-5.18 | 0.039** | 2.66 | 1.12-6.31 | 0.027** |
| Hyperlipidemia, dyslipidemia,n(%) |  |  |  |  |  |  |
| Renal dysfunction*,n(%) | 0.78 | 0.08-7.71 | 0.83 |  |  |  |
| Smoker,n(%) | 1.60 | 0.70-3.66 | 0.27 |  |  |  |
| EF,% | 1.00 | 0.96-1.04 | 0.95 |  |  |  |
| LDL;mg/dL | 0.95 | 0.57-1.60 | 0.86 |  |  |  |
| TC,mmol/L | 0.87 | 0.57-1.32 | 0.52 |  |  |  |
| LP(a),mg/L | 1.00 | 0.99-1.00 | 0.36 |  |  |  |
| Cr,μmol/L | 0.99 | 0.98-1.01 | 0.48 |  |  |  |
| UA,μmol/L | 0.99 | 0.99-1.00 | 0.53 |  |  |  |
| Story of heart failure,n(%) | 0.89 | 0.29-2.72 | 0.84 |  |  |  |
| Three-vessel coronary disease,n(%) | 0.59 | 0.27-1.31 | 0.59 |  |  |  |
| Stenosis rate of target vascular area,% | 1.03 | 0.97-1.10 | 0.29 |  |  |  |
| rotational lesion,mm | 2.13 | 1.22-3.75 | 0.008** | 2.03 | 1.16-3.59 | 0.014** |
| Rotational time,s | 1.00 | 0.99-1.01 | 0.63 |  |  |  |
| Maximum RA time of each pass,s | 1.11 | 0.88-1.41 | 0.37 |  |  |  |
| No reflow,n(%) | 0.59 | 0.06-5.54 | 0.64 |  |  |  |
| Mild complications | 0.96 | 0.35-2.63 | 0.94 |  |  |  |
| Serious complication | 0.24 | 0.03-1.99 | 0.18 |  |  |  |
| Number size of burrs | 1.54 | 0.24-9.62 | 0.65 |  |  |  |
| The maximum size of burrs | 3.20 | 0.38-26.58 | 0.28 |  |  |  |
| Target vessel | 1.19 | 0.75-1.89 | 0.47 |  |  |  |

OR, odds ratio; CI, confidence interval; AMR,Angiographic microvascular resistance;CMD,Coronary microvascular dysfunction,AMR ≥ 2.5mmHg·s/cm; EF,left ventricular ejection fraction; LDL,Low Density Lipoprotein; TC,total cholesterol; Cr,creatinine; UA,uric acid; LP(a),Lipoprotein (a); RA, rotational atherectomy; rpm,revolutions per minute; Mild complications include hematoma, dissection, etc. but blood pressure is stable; Severe complications include shock and the need for auxiliary devices such as IABP or ECMO,etc. \*:P<0.1,\*\*:P<0.05

## Discussion

This study attempted to determine the effect of RA on coronary microcirculatory function and prognosis based on AMR. The main findings were as follows: (1) RA typically resulted in an increase in post-RA microcirculatory resistance and the number of patients with CMD. (2) MACE’s incidence increased significantly among patients with CMD compared to those without CMD; (3) AMR-defined CMD was determined as an independent risk factor for the occurrence of MACE events after RA. (4) Lesion length subjected to RA and DM were independent predictors of post-RA CMD.

RA pretreatment of calcified plaques significantly increases interventional device passage and improves stent release. It is the most effective treatment tool for severe calcification.[16] However, plaque debris generated by RA treatment leads to adverse effects including microcirculatory disturbances, increasing myocardial injury, and no-reflow.[17]

IMR and CFR are relative reference standards for the assessment of microcirculation.[18, 19] IMR measurement based on pressure-temperature guide wire requires repeated low-temperature saline perfusion, which greatly limits the technological application due to repeatability and poor convenience.[20, 21] CFR has application limitations as it is highly pressure-dependent on maximum congested coronary blood flow and is easily affected by changes in hemodynamics and heart rate. [22] Invasive measurements increase the unpredictable risk of surgery.

In recent years, accumulating evidence and clinical applications of CAG-related FFR are in strong agreement with guidewire-based FFR and have been highly promoted owing to their simplicity.[23–25] Numerous subsequent studies have demonstrated the high agreement of angio-FFR-derived microcirculatory resistance with wire-based IMR with an accuracy ranging from 79.8 - 87.2%.[11, 26, 27] CAG-IMR is diagnostic of microcirculatory dysfunction and is significantly and independently associated with adverse events such as cardiovascular death and rehospitalization due to heart failure. [26, 28]

Wang’s study confirmed that approximately 45.8% of patients with RA had single or multivessel CMD. [29] In the present study, we verified that RA increases the incidence of postoperative CMD in patients, and in the present study, 30.7% of patients presented with CMD after surgery, lower compared to the percentage reported previously, which may be due to the lower proportion of chronic total occlusions (CTO) and milder lesions in patients with RA in the present study.

We found that patients presenting with CMD after surgery had longer lesion lengths that required treatment with RA. We hypothesized that most of the plaque debris produced by RA surgery is much smaller in diameter compared to normal mature erythrocytes and can be removed in microcirculation without difficulty.[30] However, when the lesion subjected to RA is longer and has a larger plaque volume, the degree of calcification is more severe, the number of RA debris and microthrombi rich in atherosclerotic particles increases, reaching or exceeding the clearance capacity of the coronary microvascular reticuloendothelial system, leading to massive amounts of debris accumulating in the microvascular network, exacerbating hypoxia, stimulating inflammation, generating embolisms and arterial spasms, and ultimately presenting with an increase in the AMR, or even formation of CMD.[31, 32] Risk factors such as high LDL, cholesterol, hypertension, stenosis area, and the longest duration of single spinning, which may be associated with coronary microvascular function, as reported previously, could not be confirmed in the present study.[26, 28, 33]

Nearly 60% of postoperative CMD cases are combined with DM, and Williams et al., showed that the higher the glycated hemoglobin in diabetic patients, the higher their coronary microcirculation resistance. This may be because of the elevated concentration of glycated hemoglobin in the blood. Its toxic effect on endothelial cells causes a weakening of the antiplatelet aggregation function and a decrease in the elasticity of blood vessels, further promoting thrombosis, as well as affecting red blood cell dysfunction, exacerbating peripheral edema, and other modalities causing microcirculatory vascular changes in the heart, kidneys, retina and other microcirculation-related organs. This ultimately decreases the microcirculatory reserve function.[34] In contrast, a trend toward higher ΔAMR was found in diabetic patients, suggesting that diabetes may amplify the elevated microcirculatory resistance due to RA, further leading to CMD.

Compared to non-CMD patients, patients with CMD showed up to four times the incidence of MACE events. There may be a significant effect of RA surgery on microvascular function, which is likely to irreversibly induce myocardial damage and reduced ventricular function, ultimately leading to significant differences in long-term prognoses.[29] However, other factors may be involved in the development of MACE, and studies by Watanabe Y, Chen, Cilia L, and others suggest that MACE incidence is similarly high in patients with RA showing renal insufficiency and low EF,[35–38] and therefore, the results need to be interpreted with caution.

This study further confirmed the effect of RA on microcirculation in patients with severe calcification after surgery. The following processes can be considered to reduce microcirculation damage during RA: 1) keeping the rotary grinding head slow near fast rewind; 2) reducing the single rotation time and complying with the operation specification of low intensity and high frequency; 3) using drugs affecting microcirculation, such as nitrate, adenosine, and glycoprotein IIb/IIIa antagonists, on a case-by-case basis; 4) active application of norepinephrine and other vasoactive drugs to avoid endothelial injury, hematoma, and other complications and reduce the damage of microcirculatory function to improve prognoses of patients.

## Limitations

The limitations of this study warrant consideration. First, this was a single-center retrospective study with an inherent bias. However, we introduced AMR to assess coronary microcirculatory function to increase the reliability of observational indexes. We used objective indexes and documented data as a reference as much as possible, and we were cautious in interpreting the results of the study. Second, although the measurement of AMR was optimized based on Murray’s law, and the agreement with wire-IMR was high, prospective studies are needed to confirm the effect of AMR on the assessment of microcirculation and prognosis. Third, we only observed the immediate and short-term effects of RA on coronary microcirculation. In future studies, we plan to determine clinical outcomes in consecutive long-term observational studies and conduct prospective studies to explore their relationship.

## Conclusions

In summary, AMR ≥2.5 mmHg-s/cm after RA was determined as an independent risk factor for poor prognosis. There exists an association between rotational length, DM, and post-RA CMD.

## Data Availability

The data that support the findings of this study are available from Yan'an Hospital of Kunming City Medical Record System but restrictions apply to the availability of these data, which were used under license for the current study, and so are not publicly available.

## Abbreviations

RA: rotational atherectomy
AMR: angiography-derived microcirculatory resistance
PCI: percutaneous coronary intervention
CFR: coronary flow reserve
CMR: coronary microvascular resistance
IMR: index of microvascular resistance
MACE: major adverse cardiovascular events

## Acknowledgment

Not applicable.

## Author Contributions

QX and XFG designed the research study. XZ, QJ, GpH, TZ and JJH performed the research. XZ and QJ analyzed and interpreted the data. XZ wrote the manuscript. All authors contributed to editorial changes in the manuscript. All authors read and approved the final manuscript. All authors have participated sufficiently in the work and agreed to be accountable for all aspects of the work.

## Ethics Approval and Consent to Participate

The study was approved by the Ethics Committee of Yan’an Hospital of Kunming City (approval number: YAXLL-AF-SC-021/01). Because data were collected retrospectively, informed consent on the use ofcoronary angiography was waived given the institutional ethics regulations with regard to observational study nature.

